# Machine learning to predict retention and viral suppression in South African HIV treatment cohorts

**DOI:** 10.1101/2021.02.03.21251100

**Authors:** M Maskew, K Sharpey-Schafer, L De Voux, J Bor, M Rennick, T Crompton, P Majuba, I Sanne, PT Pisa, J Miot

## Abstract

**Background:** To optimize South Africa’s HIV response and reach PEPFAR’s 95:95:95 targets require same day initiations and patients successfully remaining on antiretroviral therapy, and remaining virally suppressed. Much effort and resources with HIV programmes have centred on tracking and tracing for those lost to follow-up (LTFU), through various back to care strategies to ensure retention of patients. However, programmes have noted the need for targeted, data driven and predictive approaches for identifying most at risk of patients for LTFU and viral unsuppression outcomes using available phenotypic data.

**Methods:** We applied predictive machine learning algorithms to anonymised, patient-level HIV programmatic data from two districts in South Africa, January 2016 - July 2019. We developed patient risk scores for two outcomes: 1) visit attendance within 28 days of next scheduled clinic visit and 2) suppression of next HIV viral load (VL). Demographic, clinical, behavioural (e.g. visit patterns) and laboratory data were investigated in multiple models as potential predictor variables of attending next scheduled visit and VL result at next visit.

**Findings:** Longitudinal data was analysed using combinations of predictor features and classification algorithms. 445,636 Patients were included in the retention model and 363,977 in the VL model. Predictive accuracy, summarized by Area Under the Curve (AUC), ranged from 0.69 for attendance at next scheduled visit to 0.76 for VL suppression, suggesting the model correctly classified patients to whether a scheduled visit would be attended in 2 out of 3 patients and whether the VL result at next test would be suppressed in approximately 3 out of 4 patients. Key predictors for both retention & VL suppression include metrics on prior late or LTFU visits, the number of prior VL tests, the time since their last visit, the number of visits on their current regimen, the patients’ age, and their time on treatment. For retention, the number of visits at the current facility and the details of the next appointment date were also predictors. For VL suppression, other predictors included the range of thee previous VL value.

**Conclusion:** Machine learning can be used to identify HIV patients at risk for defaulting on treatment and not being virally suppressed. Predictive modelling can improve targeting of interventions through differentiated models of care, increasing cost-effectiveness and improving patient outcomes.

## Background

During 2018, South Africa was estimated to have more than seven million people living with HIV [1], representing the largest single country epidemic[2] and treatment program. In September 2016, the National Department of Health revised its treatment guidelines to extend the availability of ART to all people living with HIV, irrespective of CD4 cell count and stage of disease[3]. This policy, widely referred to as “treat all” or “universal test and treat” (UTT) holds promise to offer substantial advancements not only in the health of those living with HIV[4], [5], but also in the country’s efforts to initiate and retain 95% of all people living with HIV (PLWH) that know their status on antiretroviral therapy (ART) as part of UNAIDS’ global 95-95-95 strategy. However, implementation of a policy like UTT requires a rapid scale-up and expansion of the ART program on a country-wide level; a shift that is often challenging in communities with the most cases of HIV and the largest ARV programs globally. By expanding eligibility, UTT has eliminated “pre-ART” care for most patients[6]. In many cases, this translates into more rapid initiation of ART with fewer required clinic visits prior to dispensing drugs. In many cases, the pre-ART care cascade is compressed into a single visit with same day initiation (SDI) of treatment. Though this increases uptake of ART, retention of new patients may have suffered. Other reasons patients may become loss to follow-up (LTFU) include treatment fatigue and patient mobility. The problem of patient LTFU within the public sector in South Africa has been well documented[7]. Recent estimates indicate ART coverage levels at approximately 70% in 2019 [8]. The potential for improved health through expanded ART availability will only be realized if innovative ways to promote sustained engagement in HIV care are developed and implemented.

To optimize South Africa’s HIV response and reach targets of 95% of PLHIV tested, 95% of those on ART, and 95% of ART patients virally suppressed, numbers of patients initiating and successfully maintaining viral suppression on antiretroviral therapy must increase[9], [10]. In 2018, just 54% PLWH in South Africa were virally suppressed [11]. While much effort and resources have been focused on tracing those LTFU and returning them to care, very little prior work has successfully addressed identifying those most at risk of poor treatment outcomes while still engaged in care.

Through-out all HIV/TB programmes, being retained on treatment and being virally suppressed has been shown as the most effective prevention approach in reducing new infections. Hence the most important indicators in the last 5 years in HIV programming has been achieving retention and VL suppression targets. We aimed to determine whether machine learning and predictive modelling algorithms applied to routinely collected longitudinal HIV phenotypic and clinical outcome data in South African programmes could consistently identify predictors of the following key outcomes (i) attendance at next scheduled clinic visit and (ii) suppression of next HIV viral load (VL).

## Methods

### Data source and study population

Longitudinal data between January 2012 and July 2019 from Mpumalanga and the Free State with information on demographics, clinical and antiretroviral treatment records including outcomes, clinical visits, laboratory outcomes and other HIV key indicators were used. For both outcomes namely (i) retention and (ii) suppression of next HIV viral load, anonymised patient data who had accessed HIV care (observed in routine HIV electronic records) and, during the period 2016-2018, had at least one recorded clinic visit, were used.

For the VL model, patient records needed to have least one VL test recorded between 2016 and 2018, while in the retention model, visit data was included only where a valid next scheduled visit date was recorded (between 14-99 days after current visit). Patients without a recorded visit on a recognized three-drug highly active antiretroviral treatment regimen were excluded. Additionally, patients that could have started treatment and had tests or visits prior to 2016 (and these histories would be reflected) but needed to have an observation in the window described above were included in the analytic dataset.

We defined “entry into HIV care” as the date of the first recorded clinic visit. During the study period, national treatment guidelines indicated VL monitoring for all patients on ART with the first VL at six months after initiation and then yearly thereafter. [12]

### Outcome variable/target definitions

We defined two primary outcomes for this analysis. First, attendance at next scheduled visit was categorised according to the PEPFAR definition. A visit was deemed a “missed visit” if a patient did not attend within 28 days of their scheduled appointment i.e., they were more than 28 days late. If they missed their appointment, they were considered LTFU and not retained in care. Secondly, having a detectable HIV viral load (>400 copies/mL) at next VL test was selected as the second primary outcome. These were selected as both represent established clinical treatment outcomes and are objectively defined (diagnostically measurable reading) and thus made for a good target outcome to build confidence around the approach. In addition to the previously noted exclusion criteria, for the VL outcome, we also removed any VL observation that had a recent prior VL (within the previous 6 months) due to the high probability of concordance between these results (both VL suppressed or both VL unsuppressed). We also excluded from analysis any VL tests that followed a high VL value (>1000 copies/mL) due to the majority of these repeat tests being confirmatory tests and having a high probability of also being unsuppressed. These exclusions remove more clinically “obvious” cases to a human observer and easy for the algorithm to correctly classify, and allowed us to build a model focusing on patients who have either previously been virologically stable or not yet been tested, i.e. have the prediction focus on those most at risk of converting to the new ‘state’ of virologically unsuppressed.

### Model selection criteria

Three classification algorithms were tested, namely: Logistical Regression, Random Forest and AdaBoost. The three algorithms have different approaches to learning the input data space, and different strengths, weaknesses and applications. Logistic regression is an example of a linear classifier, and best suited separating the feature hyperspace with a linear boundary between the two classes. The random forest approach is an ensemble approach consisting of a collection of different randomly composed decision trees whose results are aggregated (through a tournament style voting process) into one best of breed result. Their random nature often limits overfitting whilst controlling error making them attractive modelling tools for complex hyperspaces with non-linear separations in classes. Like Random Forest, AdaBoost (“Adaptive Boosting”) is an ensemble set of decision trees. However, it uses boosting to evaluate the performance of sub-trees sequentially as the training develops instead of waiting to the end the way random forest does.

Both the ensemble methods are designed to predict a binary classifier based on various inputs, but do not have the linear separation limitation of logistic regression. Both the ensemble learning algorithms have the benefit of upweighting the combination of variables at certain thresholds on how they correlate with an output variable – thus often being more sensitive to minority classes or multiple sub-groups that may emerge in the data. The ensemble methods also have the ability to produce both a prediction for an unseen sample, as well as a probability rating of its prediction (which logistic regression can’t).

### Model evaluation and risk grouping

The model’s performance was evaluated in terms of ***accuracy*** (proportion of observations correctly classified by the algorithm among all observations in the unseen test set), ***sensitivity*** (the proportion of known positive outcomes in the unseen test set that are correctly identified as such by the algorithm), ***positive predictive value*** also known as *precision* (the proportion of positive outcomes predicted by the algorithm that correspond to known positive outcomes in the unseen test set) and ***specificity*** (the proportion of known negative outcomes in the unseen test set that are correctly identified as such). Next, we utilized the *area under the curve* (AUC) of a *receiver operating characteristic* (ROC) curve to evaluate the broad predictive classification performance of the model. A range of 0.5 indicates no predictive power while 1.0 indicates perfect predictive power.

Finally, we iterated on the model, by looking individually at the highest and lowest ranked patients and each of their characteristics. We manually checked for areas of obvious commonality and where the algorithm might be choosing obvious answers or erroneous assumptions. For example, early models prioritized patients who had a recent (<4 months) high VL, a situation addressed in the treatment guidelines, but resulting in the algorithm over-relying on *clinically* obvious cases. This manual validation process led to removal of these cases in the final model.

### Model building

In total, 75 predictor features (engineered variables) were investigated for inclusion in the model including features relating to patient demographics (e.g. age, gender), patient visit patterns (e.g. how many appointments experienced, how often late for appointments, days of the week, days of the month, visits at how many facilities) as well as patient testing and treatment history (e.g. current regimen, number of regimen switches) among others. Laboratory, visit and patient demographic data were linked and de-duplicated and invalid or implausible fields removed as outlined in Figure 1. This analytic set was evaluated to establish the baseline prevalence of each outcome (% of all visits that were late by >28 days and % of all VL tests that were unsuppressed). The analytic set was then randomly sampled on a 70/30 split into a training (70%) and test (30%) set. The training set for each outcome was then down-sampled to 50/50 positives (visits classified as late in the retention dataset and unsuppressed VL result in the VL dataset) and negatives (visits not classified as late and suppressed VL result suppressed) to produce a balanced set of positives and negative examples for the classification algorithm to learn from. This step also addresses bias tendency towards predicting the majority class observed in many machine learning algorithms known as the *Class Imbalance Problem* [13]. An additional model was tested with unbalanced class membership (60-40) in the training data for comparison.

**Figure 1:**
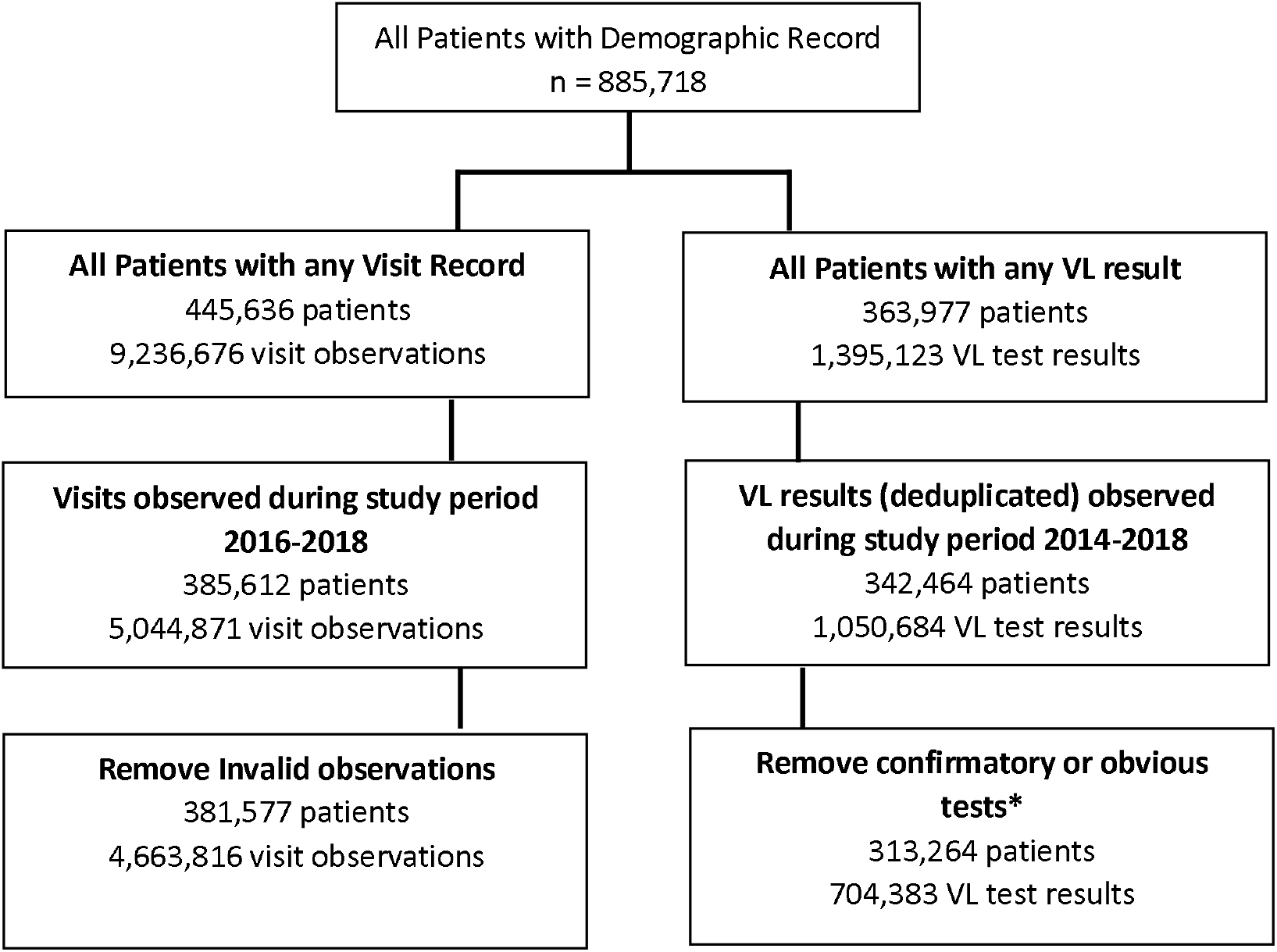
Flow chart of data source inclusion. **\*Notes:** All VL tests immediately following a high test (>=1000) and subsequent tests done < 6 months after first test removed All visits with a ‘next appointment date’ that was either missing or more than 99 days r were removed

The classifier algorithm is then trained by input of predictor features as well as the specified target outcomes to produce an optimal configuration (predictive model) such that the predictor features correspond to the specified target outcomes as often as possible. After the model was trained, we separated the unseen test set into predictor features and outcomes. The unseen predictor features are given to the model which generates its predicted outcome for each observation (in this case, observations were each scheduled visit for the retention outcome and each viral load test for the VL outcome). The predictions are then scored for accuracy against the known outcomes in the unseen test set.

### Data on predictor features

Potential predictors of retention and VL suppression were obtained from available routine clinical records. These data included patient information such as demographics, drug regimen, visit history (incl. some facility information) and schedules, and lab results.

The patient demographic, visit and laboratory data were used to create additional engineered variables or “features” that are calculated or inferred from the existing raw data. For example, the sequence of each visit of an individual patient was enumerated (1st, 2nd, etc) as well as the regimen that the patient was on at that visit, or the day of the week or day of the month that the visit happened to be scheduled for. Certain missing data was also encoded, for example, the feature “previous viral load value” was encoded with -1 for patients who had not had a previous test.

### Targets Variables

The dependent (‘target’) variable representing the outcomes of interest was encoded in the dataset for each observation. In binary classification, the target variable needs to be encoded as 1 for ‘true’ and 0 ‘false’. These states represent whether the input sample was expected to be part of the target class of ‘positives’ or not. For the VL model the target was encoded as 1 or ‘True’ if the test had a result of more than 400 VL copies/mL, and 0 or ‘False’ if the test result was less than that. For the retention model the target was calculated as whether the current visit was followed by recorded visit at most 28 days after the expected ‘next scheduled appointment date’ (Figure 4).

### Ethics statement

This study involved secondary analysis of de-identified data collected as part of routine care. Analysis was approved by the Human Research Ethics Committee of the University of the Witwatersrand (M140201).

## Results

For the purposes of the objective of the current paper and the inclusion criterion, a total of 445,636 patients were included in the retention model and 363,977 in the VL model. These patients accessing HIV care were analysed in different models using multiple combinations of input features and classification algorithms (Figure 1). Nearly one third (30%) of patients were male, median age of 39 years (IQR 31 – 49 years) at time of visit. In the retention dataset, patients had a median of 18 (IQR 10 – 25) visits since entering care and had been in care for a median of 31 (IQR 15 – 43) months. The vast majority (91%) of patients visited a single facility during the period under analysis.

### Input Features and Baselines

We generated 75 potential input features per visit and 42 features per VL test, extrapolated from the observation data of the visits, laboratory results and patient demographics. For each final model, these were reduced to a priority group of consistently predictive features as summarized in Table 1. The retention and VL prediction models were built using the AdaBoost and Random Forest [14] binary classification algorithms respectively, from the scikit-learn [15] open source project and tested against unseen data to evaluate predictive performance.

**Table 1 A:** Input Features for the LTFU Model

| ***Input Feature*** | ***Description*** |
| --- | --- |
| 3 Days Late Ratio | Ratio of patient's historical visits which they have been late by 3+ days |
| 28 Days Late Count | How many times previously has the patient been late by 28+ days |
| Visits at This Facility | How many visits has the patient had at this facility (in total) |
| Number of VL Tests | The number of VL tests the patient has had (in total) |
| Months Since First Visit | Months since the patients' earliest recorded visit in patient record |
| Months Since Last Visit | Months since the patients' most recent recorded visit in patient record |
| Current Age | Patient's age at the current visit |
| Day of Month Next Appointment | Day of the month (1-31) that the next appointment is schedule for |
| Last VL Value | Patient's last VL test value in copies/mL |
| Day of Week Next Appointment | Day of the week (1-7) that the next appointment is schedule for |
| # Visits on Current Regimen | The number of recorded sequential visits the patient has had on the current ART treatment regimen |
| Is Male | Gender of the patient encoded as 1 for Male, and 0 for Female |

**Table 1 B:** Input Features for the unsuppressed VL Model

| ***Input Feature*** | ***Description*** |
| --- | --- |
| Age Started ART | Patient's age on starting ART |
| Last VL Value | Patient's last VL test value in copies/mL |
| Duration on ART | Total number of months on ART treatment ever |
| Month of Test | Calendar Month of Test |
| #Visits on Current Regimen | The number of recorded sequential visits the patient has had on the current ART treatment regimen |
| # of Visits Ever | The number of recorded visits the patient has had ever |
| Visits Miss Ratio | The proportion of patient's historical visits which they have missed completely |
| Months Since Last VL Test | Months since a last VL test was taken |
| Months Since Last Visit | Months since the patient was last recorded to have visited a facility |
| Year of Test | The year of the test |
| # Missed Visits Ever | The number of appointments the patient has missed completely ever |

For the retention model, the test set consisted of 1,399,145 unseen visits randomly selected from across 2016-2018. The test set’s baseline (average) prevalence of missed visits was 10.5% (n=146,881 visits), consistent with the LTFU prevalence observed in both the full data set and the training set. This observed baseline was comparable with meta-studies of LTFU at 1 year in South Africa 2011-2015 [16]. For the unsuppressed VL model, the dataset was split into training and testing sets, with the test set consisting of 30% (n=211,315) of the original unseen tests randomly selected from across the study period. In the VL test set, there were 21,679 unsuppressed (>400 copies/mL) viral load results for a baseline prevalence of unsuppressed VL results of 10.3%.

### Retention Model Training Set Results

The AdaBoost classifier was trained with a 50:50 balanced sample of the modelling set which resulted in 343,078 of each visit classification (*missed* or *not missed* visits) in the training set. The retention model correctly classified 926,814 of the test set (∼1.4m visits) correctly, yielding an accuracy of 66.2% (Table 2A). In total, 89,140 patients attending their scheduled visit late or never were correctly identified out of a possible 146,881 available, yielding a recall (or sensitivity) of 60.6% all positives, and a miss rate (or false negative rate) of 39.4%. Interestingly the model’s precision of predicting a negative (or specificity) was very high at 94%, suggesting that many visits attended on time (i.e. lower risk) are simpler to recognize.

**Table 2:** Late visit model metrics based on A) balanced and B) unbalanced training sets

| A: Balanced 50:50 training set |  |  |  |  |
| --- | --- | --- | --- | --- |
|  | Specificity | Sensitivity | F1-score | # Observations available |
| Negatives | 94% |  | 0.78 | 1,252,264 |
| Positives |  | 61% | 0.27 | 146,881 |
| AUC |  | 0.688 |  | 1,399,145 |
| Accuracy |  | 66.2% |  | 1,399,145 |
| B: Unbalanced 60:40 training set |  |  |  |  |
|  | Specificity | Sensitivity | F1-score | # Observations available |
| Negatives | 92% |  | 0.87 | 1,252,366 |
| Positives |  | 41% | 0.29 | 146,779 |
| AUC |  | 0.688 |  | 1,399,145 |
| Accuracy |  | 78.6% |  | 1,399,145 |

**Notes:**
Negatives are the class of observed visits that were not followed by a missed appointment
Positives are the class of observed visits that were followed by a missed appointment
F1 score is a weighted mean of precision and recall such that 1.0 is the best score, helpful for comparing models rather than accuracy
Accuracy is the number of correct predictions out of the total number of predictions over the test set observations

The AdaBoost classifier was trained with an unbalanced 60:40 sample of the modelling set. This translated into 343,180 missed visits and 514,770 visits attended on time in the training set. The retention model correctly classified 1,100,341 of the test set (∼1.4m), yielding a higher accuracy of 78.6% (Table 2B). However, 59,739 of the late patients were correctly identified, yielding a lower recall (or *sensitivity*) of 40.6% all positives, and an increased miss rate (or *false negative rate*) of 59.3%. The model’s precision of predicting a negative (or *specificity*) remained high at 92%, further suggesting that attended scheduled visits are easier to identify than missed visits.

The two models demonstrated a trade-off in accuracy, precision and recall that can be manipulated in the training of the models. However, the predictive power or utility of the model to separate between classes – represented by the AUC metric – stayed consistent between the models. The two ROC curves are depicted in Figure 2A and Figure 2B with the same AUC, and identical shapes.

**Figure 1:**
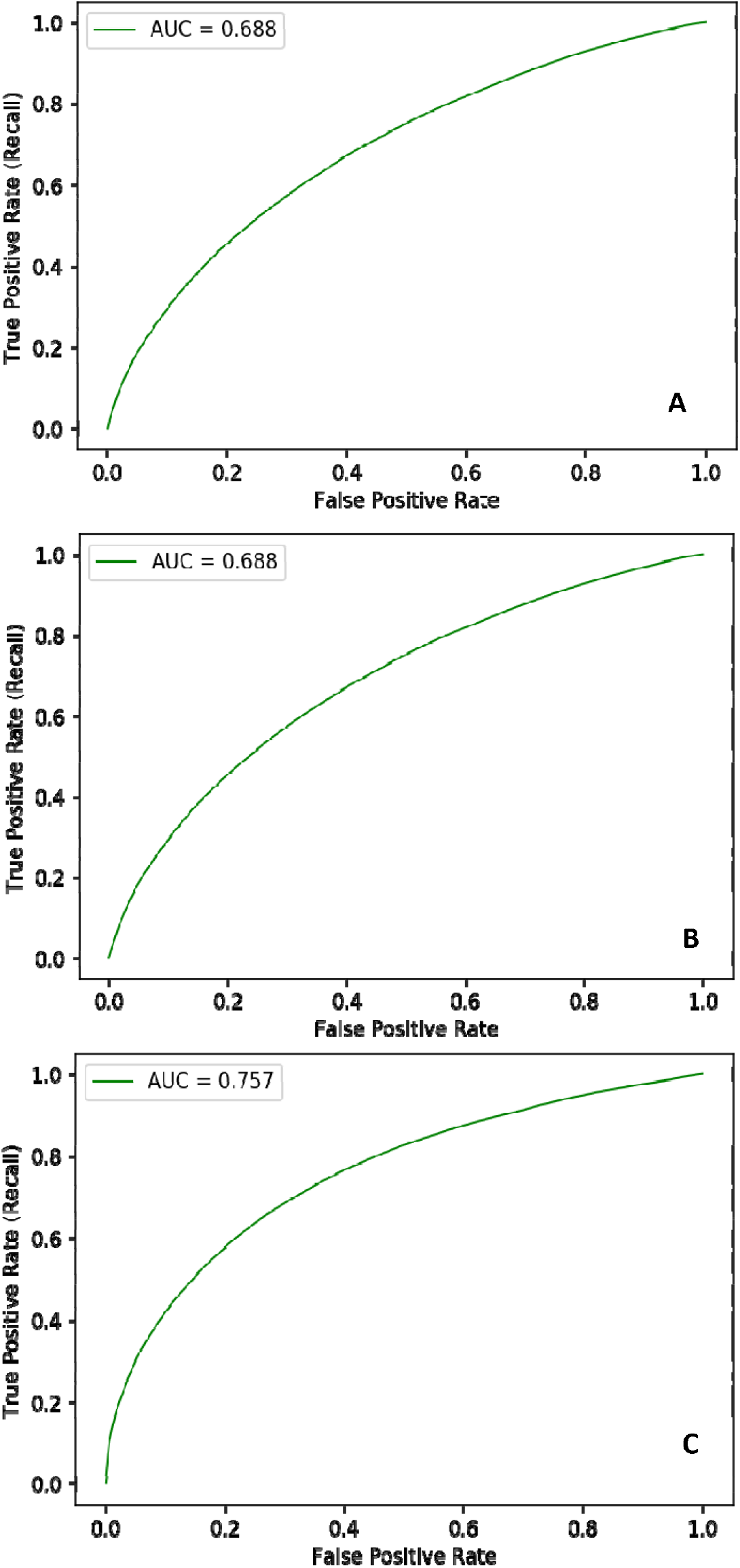
ROC Curve of A) 50:50 balanced late visit classifier, B) 60:40 unbalanced late visit Classifier and C) 50:50 balanced unsuppressed VL classifier

### Suppressed VL Model Training Set Results

For the suppressed VL model, the final training set was down-sampled to 101,976 tests, such that it had 50:50 balanced sample. The model correctly classified 153,183 VL results out of the test set of 211,315 correctly, yielding an accuracy of 72.5% (Table 3). In total, 14,225 unsuppressed viral load tests were correctly predicted out of a possible 21,679 unsuppressed test results, yielding a recall (or *true positive rate*) of 65.6% all positives, and a miss rate (or *false negative rate*) of 34.4%. The model’s specificity of predicting a negative (or *negative predictive value*) was very high at over 95%, again suggesting that suppressed VL results (i.e. lower risk) are simpler to recognize. Overall, the model had an AUC of 0.758 (Table 3, Figure 2A).

**Table 3:**
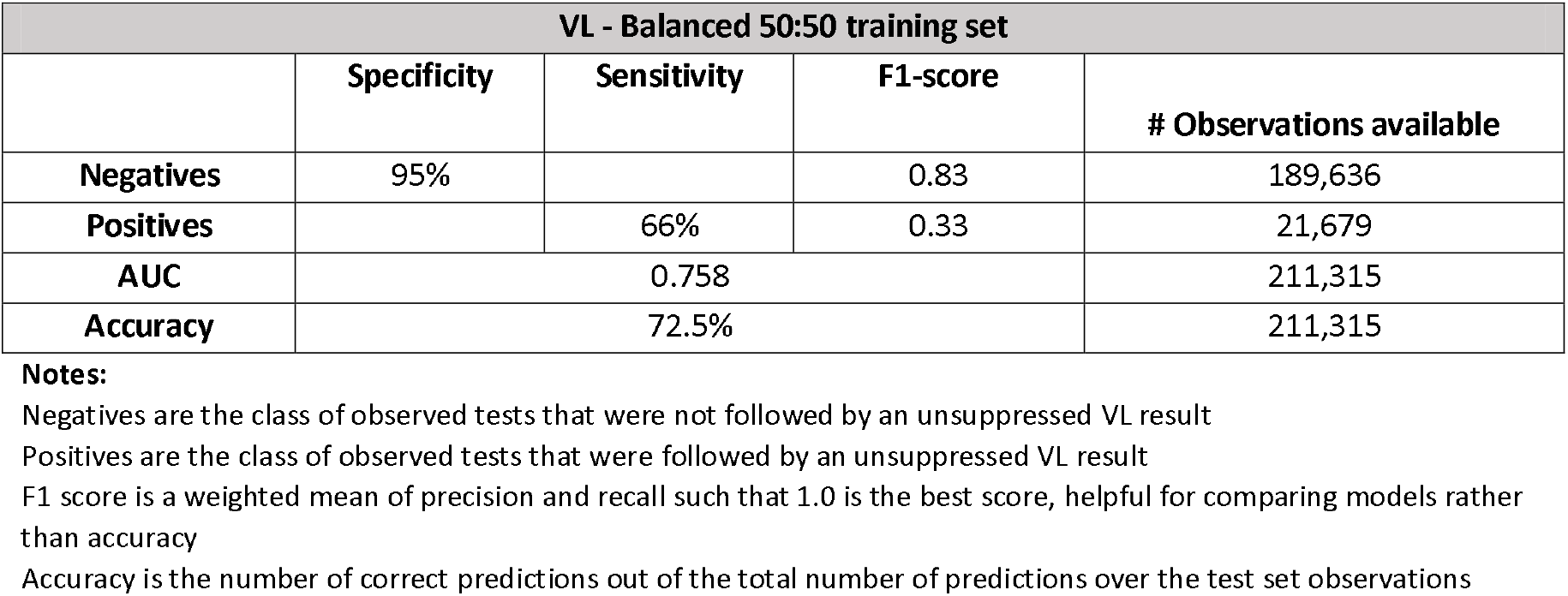
Unsuppressed VL model metrics based on balanced (50:50) training sets

| VL - Balanced 50:50 training set |  |  |  |  |
| --- | --- | --- | --- | --- |
|  | Specificity | Sensitivity | F1-score | # Observations available |
| Negatives | 95% |  | 0.83 | 189,636 |
| Positives |  | 66% | 0.33 | 21,679 |
| AUC |  | 0.758 |  | 211,315 |
| Accuracy |  | 72.5% |  | 211,315 |

**Notes:**
Negatives are the class of observed tests that were not followed by an unsuppressed VL result
Positives are the class of observed tests that were followed by an unsuppressed VL result
F1 score is a weighted mean of precision and recall such that 1.0 is the best score, helpful for comparing models rather than accuracy
Accuracy is the number of correct predictions out of the total number of predictions over the test set observations

### Feature Importance

The original set of over 75 input features for the retention model and 42 input features for the unsuppressed VL model was permuted into different groups of features and the change in predictive power (as measured by AUC) of the model for each permutation was calculated. This process retains groups of features that together improve predictive power and removes features or groups of features that contribute little or no improvement to AUC. Figures 3A and 3B illustrate their relative importance in helping *correctly and repeatedly* classify a particular observation as a correct or incorrect prediction of the target outcome. The features with higher importance help the algorithm distinguish between its classifications *more often, more correctly* than features with lower importance. For example, in the retention model (Figure 3a), gender represented in the Boolean variable ‘Is Male’ has some correlation with the missed visit target outcome and measurably more than the eliminated features that had zero correlation. However, it’s clear that the algorithm relied on correlations in the patients’ prior behaviour (frequency of lateness, time on treatment etc) to segment the risk of outcome and together these features described more of the difference than gender alone.

**Figure 3A:**
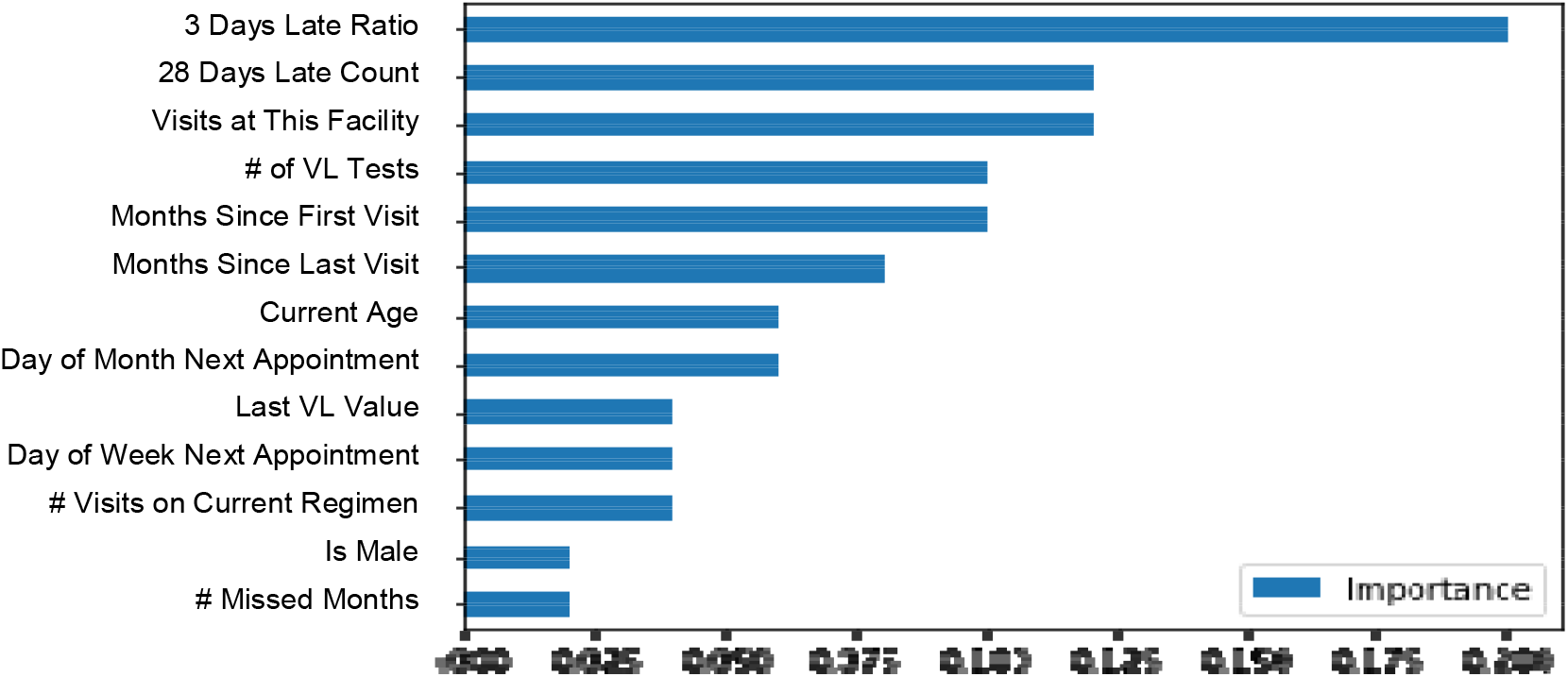
Final input features included in late visit model ranked by importance

**Figure 3B:**
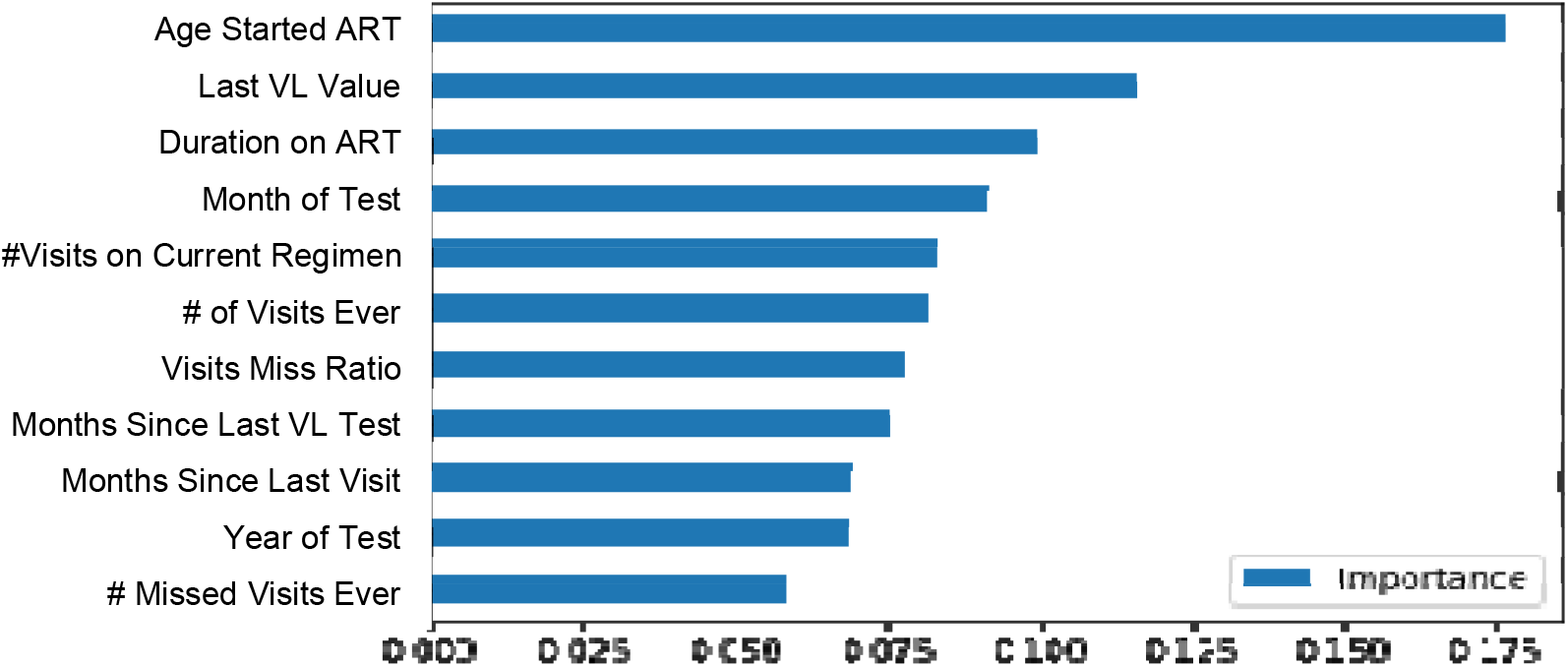
Final input features included in unsuppressed VL model ranked by importance

It was clear that prior patient behaviour and treatment history were extremely important in predicting both visit attendance and viral load result; and that traditional demographic features were less powerful predictors. These more powerful predictors can be used to further stratify populations by risk and segment more granularly for targeted interventions and differentiated care.

## Discussion

As South Africa increases efforts towards 95-95-95 goals, knowing which patients require additional services and interventions in order to achieve successful treatment outcomes at each step of the care cascade is critical. As national health monitoring systems expand to collect large volumes of increasingly detailed data, application of data science technology and methodology to these and other data sources holds the potential to improve individual health across population groups.

Our results suggest some important implications for the application of this methodology in clinical practice. First, utilising only routinely collected demographic, visit and laboratory data, the predictive models were able to correctly predict attendance in 66 -79% of scheduled clinic visits and VL suppression in 76% of viral load test results, two key features of UNAID’s 95-95-95 targets. Second, the shape of the ROC curve allows us to identify thresholds and ranges of differing group risk profiles (groups that represent an important share of the those with each outcome). For example, in the unsuppressed VL model, 20% of the population accounted for >50% of the unsuppressed VL results indicating different risk strata within population groups. Thresholds can be identified to segment the population into risk groups (“Red” = high risk of outcome, “Yellow” = medium risk of outcome and “green” = low risk of outcome). This stratification of risk groups can be leveraged for gains in predictive power estimated through population segments. Targeting at-risk patients before they become lost to follow-up or virally unsuppressed allows for greater resource use efficiencies.

Third, we demonstrated high specificity (94% & 95% for the retention and VL models, respectively) and sensitivity >60% for both models. This, combined with low positive predictive value (PPV ∼20%), indicates potentially that low risk or so-called “green” patients are readily and accurately identified by the model and have common traits. In contrast, patients at high risk of poor outcomes or so-called “red” patients can be readily identified as “not green” but demonstrated much heterogeneity in terms of presentation and underlying reason for disengaging from care. Or, to paraphrase the Anna Karenina principle, [17] *“all happy patients are alike; each unhappy patient is unhappy in its own way*”.

Our results should be interpreted in light of some limitations. First, as anonymised routinely collected facility level data was used to fit the models, it was not possible to trace missing data nor correct erroneously linked visit and laboratory data. Second, facility-level data used to define the outcome in each of the models does not account for all silent transfers to another facility and so, to the extent that this outcome misclassification has occurred, we will over-estimate LTFU and blur the predictive efforts of the model. Future models would benefit from being fit to national cohorts where a system-wide view accounts for the effect of silent transfers.[18] Third, one of the recognised limitations in the nature of black-box machine learning methods like classification trees is that whilst these features contribute to difference in risk, we can’t yet interpret or explain *how* or *why* they contribute [19]. The next step is to understand which values or ranges of the feature’s spectrum in combination with the other variables at certain levels, may correlate with a certain outcome. For example, it is unclear if the risk relationship between visits and age and poor outcomes is with younger people coming on time, or older people being late or vice versa. Additional analysis and modelling activities are underway to provide interpretable descriptions of how the algorithm is able to segment the populations of observations so well. Finally, much of the understanding as to *why* certain visit or demographic features are important may lie in more subtle social nuances related to an individual’s social circumstances and health-seeking behaviours (such as disclosure, employment or family support) [20]. To this end, our approach can be utilised and further refined by applying it to different data sources with richer social and behavioural features.

Despite these limitations, the analysis has several strengths. First, our findings highlight the importance of better understanding the risk profile of individuals, supporting recent calls for advancing data science towards precision public health models [21]. Accurately identifying those at risk for poor treatment outcomes will allow for health care services to better triage patients, improving efficiency and resource utilization. By prioritizing those most at-risk, clinics can realize better health outcomes without additional investments in data collection and staff. In addition, the results of the algorithm can also be aggregated and used to risk score population sub groups at facility level to identify where programs need to target specific interventions.

Second, while most retention interventions are directed at tracing patients disengaged from care and then returning them to care, our model offers the opportunity to shift the focus and resources directed at retention efforts to a period while the at-risk patient is still engaged at point of care. By anticipating future issues before any visible clinical signs are present (e.g. an unsuppressed VL), clinics can intervene pro-actively while patients are still accessible, engaged in health services and provide targeted services earlier. Early detection of patients at high risk of becoming virologically unsuppressed has implications not only for the individual patient’s health, but also for the risk of onward transmission of the virus and impact on breaking transmission chains. [22]–[24]

## Conclusions

A need exists to leverage predictive models and machine learning to identify and target HIV patients at risk to defaulting on treatment and not being VL suppressed. This approach should be extended to other key HIV outcomes, allowing for use of a cost-effective and precision programming approach. Additionally, by anticipating future outcomes before any visible signs and/or unwanted outcomes are present (e.g. an unsuppressed VL), specific targeted interventions can be designed on identified subsets of the treatment cohorts allowing for cost-effective differentiated models on care and treatment to be applied across the cascade.

## Data Availability

Access to primary data is subject to restrictions owing to privacy and ethics policies set by the South African Government.

## Author Contributions

Study Design: MM, KSS, LDV, TC

Data collection: TC

Data analysis: KSS, LDV

Funding acquisition: JM

Data Interpretation: KSS, LDV, MM, TC

Supervision: MM

Validation: KSS, LDV

Writing – original draft: Mhairi Maskew

Writing – review and editing: All authors

